# Social capital and psychological distress during Colombian coronavirus disease lockdown

**DOI:** 10.1101/2020.09.04.20187914

**Authors:** Carmen Cecilia Caballero-Domínguez, Jeimmy De Luque-Salcedo, Adalberto Campo-Arias

**Author notes:** **Corresponding author**: Adalberto Campo-Arias, Universidad del Magdalena, Carrera 32 No. 22-08, Bloque 8, Gorgona, Segundo piso, oficina Psicología de la Salud y Psiquiatría, Santa Marta, Colombia (Código postal 470004) Teléfono: 57 5 4381000 Ext. 3059.

## Abstract

**Introduction:** This study aims to establish the association between low capital social (CS) with some indicators of psychological distress.

**Methods:** A cross-sectional study was carried out using an online questionnaire that evaluated demographic variables, social capital, coronavirus disease perceived stress, depression risk, insomnia risk, and suicide risk. SC was taken as an independent variable, and symptoms indicating psychological distress was handed as dependent variables. Odds ratios (OR) were established with 95% confidence intervals (CI), using binary logistic regression analysis.

**Results:** A group of 700 adults participated in the survey; they were aged between 18 and 76 years (M = 37, SD = 13). Low SC was associated with depression risk (OR = 2.00, 95%CI 1.34-2.97), elevated suicide risk (OR = 2.62, 95%CI 1.40-4.91) high perceived stress related to coronavirus disease (OR = 2.08, 95%CI 1.15-3.76), and insomnia risk (OR = 2.42, 95%CI 1.69-3.47).

**Conclusions:** Low CS was associated with indicators of psychological distress represented in depression risk, elevated suicide risk, high perceived stress related to coronavirus disease and insomnia risk. SC is a community social resource that could help mitigate the impact of the coronavirus disease quarantine amidst the Colombian population’s psychological health. It is necessary to deepen the SC role in psychological well-being during and after the coronavirus disease epidemic.

## 1. Introduction

In recent years, health research has gone from traditional epidemiology to considering the social determinants of health (Reinenger *et al*., 2013). The collective attributes and family norms that occur in the dynamics of interactions between individuals and society, characterized by reciprocity and social action, and increase well-being (Bravo-Vallejos, 2017). From this perspective, social capital (SC) is considered as a social determinant of public health that can explain some inequities and inequalities observed in the health sector (Bravo-Vallejos, 2017).

In 1893, Durkheim proposed SC’s concept, which can be defined as the ability to access resources by maintaining stable formal and informal relationships based on trust, mutual aid, and cooperation (Flores and Rello, 2003). Furthermore, SC is a social resource that favours joint and decisive action during crises, the deployment of adaptive responses to stressful situations, promoting psychological well-being, both for individuals and the collective (Aldrich, 2011).

SC is a construct that comprises two components that complement each other, cognitive social capital (CSC) and structural social capital (SSC). The SSC includes the set of perceptions, values, norms, shared beliefs of the social organization, and trust in other members of the community (Brune and Bossert, 2009). Furthermore, SSC is the behavioural expression of SC and refers to observable and contextual components capable of linking individuals and groups (Aldrich, 2011). SSC includes vertical or bond share capital and horizontal share capital. The vertical SSC reflects the power relations established between subjects or groups with institutions for the management of social, economic, and political resources (Kripper and Sapag, 2000). Meanwhile, the horizontal SSC covers relationships between peer groups, either through close networks such as family, neighbours, friends (union SSC) or between acquaintances or more heterogeneous groups (approach SSC) (Islam *et al*., 2006).

Previous studies indicate the theoretical link between SC and psychological health, healthy behaviours, access to information, health education, medical services, psychological care, and psychosocial support (Bravo-Vallejos, 2017). Some authors observed that CSC (trust and reciprocity) is more associated with psychological health problems than SSC (Poblete *et al*., 2008), such as the risk of major depression, suicidal ideation, perceived stress, and risk of insomnia.

To date, research has focused on the association between SC and major depressive disorder. For example, in King County, Washington, in 356 Mexican women, low social capital was found to be associated with depression (Bassett and Moore, 2013). In China, in 928 older adults, trust, reciprocity, and social participation were related to less depression (Cao *et al*., 2014). In Brazil, in 15,052 participants, it was found that lower SC was associated with a higher probability of presenting symptoms of depression (Paiva *et al*., 2020).

A smaller number of studies indicate the link between low SC with other psychological health problems. In Okayama, Japan, 10,094 older adults in rural areas showed that perceptions of mistrust and little reciprocity at the community level were associated with suicidal ideation (Noguchi *et al*., 2017). Similarly, in 11 European countries, in 22,227 participants was found that greater trust was associated with a lower suicide rate (Kelly *et al*., 2009). In Brazil, 1,180 participants, SC was directly associated with sleep quality (Muñoz-Pareja *et al*., 2016).

The CSC has been associated with psychological health and a higher capacity to respond and recover effectively from traumatic events such as armed conflicts, natural disasters, terrorist attacks, epidemics, or confinement (Cardozo *et al*., 2017; Xiao *et al*., 2020). In the present coronavirus disease pandemic (COVID-19), confinement is a stressor for physical and psychological health (Xiao *et al*., 2020). The physical isolation, uncertainty about a vaccine, COVID-19 contagion risk (Zandifar and Badrfam, 2020), and disinformation generate individual and collective concerns (Bo et al., 2020). Lack of cooperation networks and establishing relationships with the community based on mistrust and little social support limit the coping strategies of this stressor, increasing the possibility of psychological problems (Aldrich, 2011). In China, in 170 participants, low SC was associated with high stress and insomnia during confinement due to the COVID-19 (Xiao *et al*., 2020).

This study addresses the relationship between CSC and some symptoms indicative of psychological distress: depression risk, elevated suicide risk, high perceived stress related to COVID-19, and insomnia risk. To date, in Latin America, no studies show the association between SC and psychological distress (Kripper and Sapag, 2000). It is essential to study SC during confinement due to COVID-19 and to know its relationship with psychological distress. This information is fundamental for the design of practical measures that reduce the psychological impact of preventive isolation at the community level, beyond the reductionist approach of individual risk factors.

The objective of this study was to estimate the association between CSC and some indicators of psychological distress during confinement due to the COVID-19 epidemic in the general Colombian population.

## 2. Methods

### 2.1 Design and ethical issues

An analytical cross-sectional observational study was performed. It was were respected the ethical principles promulgated in the Resolution 008430 of 1993 of the Colombian Ministry of Health and Declaration of Helsinki. An institutional research ethics committee approved the project. All participants must agree to participate.

### 2.2 Population and sample

A non-probability snowball sampling was implemented. It was estimated a sample of at least 384 participants, adequate for a prevalence of 50% for the study variable, and margin of error of 5% (Hernández, 2006). This sample is indicated to explore associations between variables with sufficiently narrow confidence intervals.

### 2.3 Instruments

#### 2.3.1 Cognitive Social Capital Scale (CSCS)

The CSCS measures global social capital and explores social cohesion and trust in the neighbourhood and nearby community, using five questions with polytomous response options that are rated from 0 to 3. The previous study did not report internal consistency (Martin *et al*., 2004). In the present investigation, scores less than or equal to five were categorized as low SC. The CSCS showed Cronbach’s alpha of 0.79.

#### 2.3.2 Depression risk

The depression risk was quantified with the Well-Being Index (WBI) measures psychological well-being, a positive way to report depressive symptoms. It consists of five items with Likert-type response options from 0 to 3. The total score can be from 0 to 15; it was established as a cut-off point for the risk of depressive episode score equal to or less than nine (World Health Organization, 1998). In the Colombian context, the scale showed adequate performance in a previous study (Campo-Arias *et al*., 2015). In the present study, the WBI presented Cronbach’s alpha 0.86.

#### 2.3.3 Suicide risk

The suicide risk was measured with the Suicidal Ideation Scale of the Center for Epidemiological Studies in Depression (CES-D-SI). The CES-D-SI consists of 4 items that quantify the presence of elevated suicide risk during the last two weeks. The CES-D-SI presents five response options ranging from 0, 1-4, 5-8, 9-11, and 12-14 days (Roberts, 1980). The scale in a previous study in Colombia showed high satisfactory consistency of 0.86. The total score can be between 5 and 20 (Pineda-Roa *et al*., 2018). In the present study, scores equal to or greater than nine were categorized as an elevated suicide risk. In the current study, the CES-D-SI showed Cronbach’s alpha 0.63.

#### 2.3.4 Perceived Stress related to COVID

The perceived stress was evaluated with the Perceived Stress Scale for COVID (PSS-C). The PSS-C provides a measure of stress related to the COVID-19 epidemic and assesses the overall psychological response to stressful situations. It consists of 10 items with Likert-type response options from 0 to 4. Scores higher 24 indicate high perceived stress related to COVID-19. This scale in a previous study during the epidemic in Colombia showed a satisfactory Cronbach’s alpha (Pedrozo-Pupo *et al*., 2020). The PSS-C showed Cronbach’s alpha of 0.88 in this sample.

#### 2.3.5 Insomnia risk

The insomnia risk was explored with the Athens Insomnia Scale (AIS). The AIS is a tool to assess insomnia risk. It has eight items that are rated from 0 to 3. The higher the score, the higher the risk of insomnia (Soldatos *et al*., 2000). The AIS showed high consistency in a previous Colombian study (Campo-Arias *et al*., 2020). The cut-off point for insomnia risk was ten or more, suggested for the non-clinical population (Soldatos *et al*., 2000). The AIS showed Cronbach’s alpha of 0.87 in the present sample.

### 2.4 Procedure

An online questionnaire was implemented to evaluate SC and symptoms indicating psychological distress in the Colombian population. The questionnaire was shared using different social networks, email, and WhatsApp to the researchers’ contacts. The questionnaire ran from March 30 to April 8, 2020, two weeks after the first COVID-19 infections and the start of the quarantine that the Colombian government declared.

### 2.5 Statistical analysis

The reliability of the applied scales was initially estimated using Cronbach’s alpha coefficients. Variables were dichotomized for bivariate analysis. The SC was taken as the independent variable and the risk of major depression, elevated suicide risk, insomnia risk, and high perceived stress related to COVID-19 as dependent variables. Odds ratios (OR) were established with 95% confidence intervals (CI). The associations were adjusted according to Greenland’s recommendations (1989) using logistic regression. The Hosmer-Lemeshow goodness of fit test was computed for each adjustment (Hosmer *et al*., 1991). Statistical analysis was performed with IBM-SPSS version 23.0.

## 3. Results

A total of 714 people completed the questionnaire, of which 14 (2.0%) were eliminated because they were people residing abroad at the time of the pandemic. Ages were observed between 18 and 76 years (M = 37.1, SD = 12.7). Table 1 expands the demographic information and the indicator of psychological distress.

**Table 1.** Frequency of demographic variables and indicators of psychological distress.

| **Variable** | **n** | **%** |
| --- | --- | --- |
| **Age (years)** |  |  |
| Between 18 and 29 | 248 | 35.4 |
| Older than 29 | 452 | 64.6 |
| **Marital status** |  |  |
| Single, separated or widowed | 364 | 52.0 |
| Free union or married | 336 | 48.0 |
| **Gender** |  |  |
| Female | 476 | 68.0 |
| Male | 224 | 32.0 |
| **Academic achievement** |  |  |
| High school or less | 72 | 10.3 |
| College or more | 628 | 90.7 |
| **Socioeconomic status** |  |  |
| Low or low middle | 420 | 60.0 |
| High middle or high | 280 | 40.0 |
| **Elevated suicide risk** |  |  |
| Yes | 53 | 7.6 |
| No | 647 | 92.3 |
| **High perceived stress related to COVID-19** |  |  |
| Yes | 53 | 7.6 |
| No | 647 | 92.3 |
| **Depression risk** |  |  |
| Yes | 428 | 61.1 |
| No | 272 | 38.9 |
| **Insomnia risk** |  |  |
| Yes | 294 | 42.0 |
| No | 406 | 58.0 |
| **Low social capital** |  |  |
| Yes | 168 | 24.0 |
| No | 532 | 76.0 |

In the bivariate analysis, low SC was significantly associated with depression risk, elevated suicide risk, high perceived stress related to COVID-19, and insomnia risk. All associations remained significant after adjusting for sex and marital status (see Table 2).

**Table 2.** Associations between low CSC and indicators of psychological distress.

| Variable | Crude OR<br>(95%CI) | Adjusted OR<br>(95%CI)<sup>a</sup> |
| --- | --- | --- |
| Depression risk | 2.19 (1.49-3.23) | 2.00 (1.34-2.97)<sup>1</sup> |
| Elevated suicide risk | 2.86 (1.53-5.32) | 2.62 (1.40-4.91)<sup>2</sup> |
| High perceived stress related to COVID-19 | 2.23 (1.24-3.98) | 2.08 (1.15-3.76)<sup>3</sup> |
| Insomnia risk | 2.56 (1.79-3.65) | 2.42 (1.69-3.47)<sup>4</sup> |

<sup>a</sup>By gender and marital status.
<sup>1</sup>Hosmer-Lemeshow's test: $X^2 = 3.34$ , $df = 4$ , $p = 0.502$ .
<sup>2</sup>Hosmer-Lemeshow's test: $X^2 = 1.26$ , $df = 4$ , $p = 0.867$ .
<sup>3</sup>Hosmer-Lemeshow's test: $X^2 = 0.21$ , $df = 4$ , $p = 0.995$ .
<sup>4</sup>Hosmer-Lemeshow's test: $X^2 = 3.31$ , $df = 5$ , $p = 0.651$ .

## 4. Discussion

In the present investigation, low SC was associated with indicators of psychological distress, depression risk, elevated suicide risk, high perceived stress related to COVID-19, and insomnia risk in a Colombian sample during confinement due to the COVID-19 epidemic.

In the present study, an association was found between SC and depression risk. This finding is similar in precedents during regular times that described an association between SC and depression in 356 women in the United States (OR = 1.46, 95%CI 1.14-1.88) (Bassett and Moore, 2013), and in 928 older adults in China, in whom it was observed that trust (β = −0.137, p < 0.001), reciprocity (β = −0.166, p < 0.001) and social participation (β = −0.137, p = 0.008) was related to lower scores in depression (Cao *et al*., 2014). Consistent with the above, low SC was associated with depressive symptoms (OR = 2.07, 95%CI 1.57-2.72) in 15,052 adults in Brazil (Paiva *et al*., 2020). Less cohesive communities and neighbourhoods with little reciprocity among members offer less support to isolated people, thereby increasing the risk of depression (Flores and Rello, 2003).

This research low SC was associated with elevated suicide risk. Before the pandemic, mistrust and little reciprocity in the community context were reported to be associated with suicidal ideation (OR = 1.98, 95%CI 1.02-3.81) in Japanese people (Noguchi *et al*., 2017); consistently with the previous findings, higher trust was related to lower suicide rate (β = −2.77, 95%CI –4.89- −0.65) in the European population (Kelly *et al*., 2009). Low SC is associated with fewer expressions of solidarity and trust (Paiva *et al*., 2020), and it can reduce social integration and connection that give the individual a sense of life, that is, increase despair (Berkman and Seeman, 2000; Goodman *et al*., 2017). Hopelessness is a single symptom most related to suicide risk (Noguchi *et al*., 2017).

In the present study, SC was related to high perceived stress related to COVID-19. This finding is consistent with that described in China during the current COVID-19 epidemic, an association between CS with high stress was observed in 170 participants (β = 0.159, p < 0.001) (Xiao *et al*., 2020). Low CS implies reduced access to community resources during crises (Noguchi *et al*., 2017), adequate information, and lack of support to resolve daily needs and economic difficulties. This negative context favours the manifestations of acute stress (Xiao *et al*., 2020).

In the present study, low CS was associated with insomnia risk. This result is consistent with studies during epidemics and at regular times. In China, during the current confinement, insomnia was reported to be indirectly associated with CS (β = 0.488, p < 0.001) (Xiao *et al*., 2020). Furthermore, in Brazil at regular times, a direct association was found between SC and sleep quality (OR = 1.36, 95%CI 1.02-1.83) (Paiva et al., 2020). Low SC could favour the feeling of insecurity, helplessness, unpredictability, and uncertainty (Xiao *et al*., 2020). In this sense, low SC could enhance insomnia risk, directly or indirectly, as measured by other psychological problems such as anxiety, stress, or depression. Insomnia can be an isolated symptom or part of another mental disorder (Cardozo *et al*., 2017).

From the perspective of the social determinants of mental health, SC is recognized as a resource that fosters social relationships (trustworthy and reciprocal) and the deployment of adaptive responses to stressful situations (Reinenger *et al*., 2013). Cohesive community settings during a crisis, such as confinement due to the COVID-19 epidemic, promote psychological well-being, adaptation, and recovery of health (Flores *et al*., 2018). Likewise, it strengthens the capacity to access health services and promote the common good and self-care behaviours, which mitigates the risk of individual and community infection (Brune and Bossert, 2009).

### 4.1 Strengths and limitations

This study is an approach to understanding the resources and attributes of the groups that characterize the CSC and its link to indicators of psychological distress, necessary for the design of actions based on community rehabilitation that promote risk mitigation strategies for contagion by COVID-19 and self-care behaviour that consolidate communities with healthy practices adapted to social and cultural contexts to overcome confinement crises. This research has some methodological limitations. Firstly, the sample is not representative of the population since sampling does not guarantee randomness; therefore, the results are not generalizable (Hernández, 2006).

### 4.2 Conclusions

It is concluded that low SC was associated with less psychological distress in this sample of residents in Colombia. Therefore, strategies aimed at strengthening SC could have a beneficial effect on the psychological well-being of the population during periods of crisis.

## Data Availability

Data will be shared with the researchers who formally request it from the author responsible for the correspondence.

## Notes

**Funding:** University of Magdalena, Santa Marta, Colombia.

### Competing Interest Statement

The authors have declared no competing interest.

### Funding Statement

The University of Magdalena and Colciencias, Colombia, funded this research (Jeimmy De Luque_Resolution 0075/2020 of Vice-Rectory of Research, University of Magdalena).

### Author Declarations

Research Ethics Board of the University of Magdalena approved this project.

